# Two-dimensional maps to predict the neurological recovery after cardiac arrest

**DOI:** 10.1101/2022.11.20.22282538

**Authors:** Aymeric Floyrac, Adrien Doumergue, Stéphane Legriel, Nicolas Deye, Bruno Megarbane, Alexandra Richard, Elodie Meppiel, Sana Masmoudi, Pierre Lozeron, Eric Vicaut, Nathalie Kubis, David Holcman

## Abstract

**Background:** Severity of neuronal damage in comatose patients following anoxic brain injury is assessed through a multimodal evaluation. However, predicting the return to full consciousness of hospitalized post-anoxic comatose patients remains challenging.

**Methods:** We present here a method to predict the return to consciousness and good neurological outcome based on the analysis of responses to auditory periodic stimulations to auditory evoked potentials. We extracted several EEG features from the time series responses in a window of few hundreds of milliseconds from the standard and deviant auditory stimulations that we considered independently. By combining these features, we built a two-dimensional map to evaluate possible group clustering. Using Gaussian, K-neighbourhood and SVM classifiers, we could predict the neurological outcome of post-anoxic comatose patients, the validity of the method being tested by a cross-validation procedure. This method was developed using data acquired retrospectively in a cohort of 29 post-cardiac arrest comatose patients, recorded between day 3 and day 6 following admission. Data from event-related potentials (ERPs) were recorded non-invasively with four surface cranial electrodes at electro-encephalography (EEG), that we computed secondarily.

**Results:** Analysis in two-dimensions of the present data revealed two separated clusters of patients with good versus bad neurological outcome. When favouring the highest specificity of our mathematical algorithms (0.91), we found a sensitivity of 0.83 and an accuracy of 0.90, maintained when calculation was performed using data from only one central electrode.

**To conclude:** statistics of standard and deviant responses considered separately provide complementary and confirmatory predictions of the outcome of anoxic comatose patients, better assessed when combining these features on a two-dimensional statistical map. The benefit of this method compared to classical EEG and ERP predictors should be tested in a large prospective cohort. If validated, this method could provide an alternative tool to intensivists, to better evaluate neurological outcome and improve patient management, without neurophysiologist assistance.

## Introduction

Sudden death by cardiac arrest (CA) is a major public health issue, affecting 55 patients out of 100,000 with nearly 40,000 cases per year in France (SFAR, SRLF, 2007). Five to 30% of the patients resuscitated after CA are alive at one year (Carr *et al*., 2009; Pell, 2003). Improvement of critical care management after CA has increased patients’ survival. Favourable outcome after discharge relies mainly on the prognostic value of brain injury that outweighs the combined effects of all other terminal organ failures (Roberts *et al*., 2013; Rossetti *et al*., 2016).

Assessment of neurological damage is usually performed 48-72 hours after CA and, optimally, after interruption of sedative drugs (Nolan et al., 2021). The evaluation is multimodal and combines, according to available local resources, clinical evaluation (Glasgow Coma Scale, photomotor and pupillary reflexes), biological markers of neural cell necrosis (NSE and S100bêta proteins), cerebral Magnetic Resonance Imaging and electrophysiological studies including electroencephalography (EEG), somatosensory evoked potentials (SSEP) and auditory evoked potentials (AEP). EEG allows the grading of post-anoxic encephalopathy as a grade 5 EEG pattern (Synek, 1988) or “highly malignant” EEG pattern (Westhall et al., 2016; André-Obadia et al., 2018), being associated with the least favourable prognosis. EEG reactivity presents additional value to predict good outcome, but given the large inter-rater variability, there is a need to develop new quantitative methods (Admiraal et al., 2020; Duez et al., 2018). The absence of cortical N20 response at SSEP after stimulation of median nerves has an almost 100% specificity for non-awakening prediction (Sandroni et al., 2014). The presence of a “mismatch negativity” (MMN), an endogenous long latency negative potential at AEP (Rohaut et al., 2009) would rather indicate a good prognosis (Fischer et al., 2006; Pfeiffer et al., 2017). These last authors found that an improved auditory discrimination over the two first days of coma predicted a good outcome, in patients treated with targeted temperature management at 36°C. By contrast, the presence of N20 or the absence of MMN does not allow predicting good or bad prognosis. Moreover, electrophysiological recordings are not always available in the intensive care unit (ICU), and may be particularly difficult to acquire at the acute phase where patients combine aggressive care (extracorporeal membrane oxygenation (ECMO), haemodialysis, mechanical ventilation), and invasive methods of monitoring, generating artefacts. At last, potentials amplitudes are smaller under sedation and more difficult to extract from the background (Yppärilä et al., 2004). Multimodal approaches combining several prognostic factors of post-anoxic coma have been proposed (Fischer et al., 2006; Bassetti et al., 1996; Oddo and Rossetti, 2014; Kim et al., 2012) but the choice of these approaches has not yet succeeded to lead to automatic and predictive analyses.

The AEP technique consists of recording cortical potentials in response to auditory stimulation delivered by earphones, using electrodes placed on the scalp. The mismatch negativity (MMN or N200), is a negative event-related potential (ERP) that occurs between 100 and 250 ms predominantly over the frontocentral scalp area. It is obtained by the susbstraction of oddball auditory stimuli (called deviant stimuli) randomly intermixed with repetitive frequent auditory stimuli also called standard or non-deviant stimuli. Deviant stimuli are distinguished by one or several features (frequency, intensity, duration…) (Fischer et al., 1999; Chausson et al., 2008; Comanducci et al., 2020). Thus, MMN reflects the ability to detect automatic auditory violations, but sensitivity to predict awakening is low (56%) with a high 93% specificity (Naccache et al., 2005). Because of lack of sensitivity in the ICU when interpreted only by visual analysis (present or absent) (Azabou et al., 2018), complementary statistical methods have been developed to analyse MMN more accurately, increasing thus the positive predictive value for awakening (Pfeiffer et al., 2017), at the cost of extension of the time of interpretation.

Taking advantage of the considerable amount of information obtained at AEP, we aimed to develop here a computational approach based on EEG features arising from the distribution of the ERP fluctuations responses during the 20 min-recording, rather than to interpret the MMN as a binary response. We used data already acquired from a homogeneous cohort of patients admitted in the intensive care unit after cardiac arrest and who all had EEG, SSEP and AEP recordings within 6 days after admission. We identified specific features from AEP, considering responses to standard and deviant auditory stimulations independently. Using a step-by-step data processing, we finally reported combined features in a two-dimensional map: we observed that patients were clustered into two groups corresponding to a different outcome at discharge whether they were able to follow verbal command or not. We then estimated the probability for a patient to be classified into one of the two groups at the acute phase.

## Patients and methods

### Procedure

This study is a retrospective single-center study performed in 29 consecutive patients between January 2014 and March 2016, successfully resuscitated after CA, with persistent coma between the 3^rd^ day and 6^th^ day following admission in the Department of Medical and Toxicological Critical Care in Lariboisière Hospital (Paris), and who completed EEG, SSEP and AEP recordings. From AEP recordings, we extracted individual features, and using a novel analysis method, we aimed to classify patients into non communicating or dead patients and communicating patients at discharge.

This study is an ancillary study of the PHRC CAPACITY AOR10109 and was approved by the ethics committee (Comité de Protection des Personnes, CPP Paris IV #2012/22). As this AEP processing was performed secondarily, physicians who were in charge of the patients could not have access to these data. Withdrawal of life-sustaining therapies was performed according to the usual guidelines (Société de réanimation de langue française, 2010).

### Clinical data

Cardiac arrest characteristics, in-hospital management and outcome data were collected according to Utstein method by the intensivists in charge during hospitalization (Perkins et al., 2015). During ICU stay, the following data were collected: age, sex, past medical history; presumed etiology categorized into non-cardiac, cardiac and undetermined; shockable rhythm; time from collapse (CA) to return of spontaneous circulation (ROSC) dichotomized into ≤ 25 or >25 minutes (Oddo et al., 2008); interval from the time of collapse (presumed time of cardiac arrest) to basic and/or advanced life support, defined as no-flow duration, and the interval from the beginning of life support until the return of spontaneous circulation or termination of resuscitative efforts, termed low-flow duration; hypothermia; Glasgow Coma Scale (GCS) on admission; SAPS II (Simplified Acute Physiology Score) (Le Gall et al., 2005); sedation.

To evaluate neurological prognosis, verbal following on command was used. Good neurological prognosis was defined by appropriate response to verbal command. Moreover, the Glasgow Outcome Scale Extended (GOS-E) was retrospectively collected at 3-6 months, when information was available.

### Electrophysiological data

We used electrophysiological data acquired between day 3 and day 6 following admission, in order not to include patients with early predictable death. However, most of them had previous EEG recording in the first 48 hours. All data were analysed or double-checked by specialists in clinical neurophysiology with at least 10 years’ experience.

#### EEG

Digital electroencephalography (EEG) recordings were performed for at least 20 min, with 21 scalp electrodes positioned according to the standard 10-20 system placement, reformatted to both bipolar and off-head referential montages, with filter settings at 0.3 Hz and 70 Hz. Repetitive bilateral auditory and painful stimulations were systematically performed. EEG was classified according to five major grades based on Synek’s classification (Synek, 1988).

#### Somatosensory evoked potentials

Median nerves were stimulated at the wrist to an intensity of 4-5 mA, greater than that needed to evoke a muscular response, and in the case of the use of neuromuscular blocking, the ERB potential amplitude was used to estimate the intensity of the stimulation. Pulse duration was 0.2 ms and stimulus rate 3 Hz. Active electrodes were placed at Erb’s point and C3 and C4 points. At least two repetitions (averages of 300 responses) were performed to assess the reproducibility of the waveforms. N20 cortical response was dichotomized into absent or present.

#### Mismatch negativity

The auditory event related potentials were elicited using the classical odd-ball paradigm technique as already described (Fischer et al., 1999). Acoustic stimuli were delivered through earphones binaurally using a randomly intertwined sequence of standard and deviant stimuli in the proportion of 86% and 14%, respectively. Event-related potentials were recorded with active electrode positioned at Fz, Cz, C3, C4 according to the International 10-20 system, reference electrode at the mastoid and ground reference at the forehead. Each recording was performed during 20 min.

Presence/absence of MMN defined as the negative peak obtained between the difference between deviant and standard response occurring in the 100-300ms time interval following stimulation. In our experience, MMN is delayed in those critically ill and sedated patients, which explains this relatively wide time window.

#### Electrophysiological analysis

All data were analyzed by at least two different neurophysiologists, blind to the neurological outcome of the patients. When artefacts were too numerous leading to unreliable conclusion, data were not considered.

### Statistical analyses for demographic and clinical data

In each group (good or bad neurological outcome), results of clinical and neurophysiological examinations were expressed as mean**±**SD [min-max] and median [IQR 25-75], when appropriate. Statistical analyses were performed with Prism 5 software (Prism 5.03, GraphPad, San Diego, USA). Comparison of frequencies in each group was analysed by the Fisher’s exact test. A value of *p*<0.05 was considered statistically significant.

### Signal processing, features identification and classification

This section is divided in three parts: 1-Signal processing, 2-Feature identification and 3-Classification using a two-dimensional map. Briefly,, we have considered specific features in a 1s duration window, then we considered a shorter window of 500ms, which contains most of the relevant features. We have then limited our analysis to this shorter window. We then split the 20 minutes recordings into two sets (two consecutive sequences of 10 min), to explore a possible adaptation between the first part of the acquisition and the last part, that could differentiate patients with good versus bad prognosis. We have then introduced two parameters to that possible adaptation analysis: a- the variance of the signal computed over 10 min and b-the correlation between the two parts of the signal.

The data corresponding to the responses obtained from standard and deviant auditory stimulations were considered independently, regardless of the mismatch negativity that was not considered here, and mathematical processing was applied as for any signal, independently of its potential significance. We identified specific features for the standard stimulations (standard deviation computed over the entire sample of 20 min and similarity) and for the deviant stimulations (number of extremum N_E and oscillation|ΔV|). At last, we combined them into a two-dimensional map, and patients formed two clusters according to their outcome. All these steps were determined without *a priori* knowledge of the patient’s prognosis.

#### 1- Signal processing

AEP obtained with standard and deviant auditory stimulations were exported in the European Data Format (EDF), which is a simple and flexible format for storage of multichannel biological and physical signals, then anonymized through a specific software we designed. Analyses were performed on all four electrodes then on one single Cz (central) electrode. To quantify the auditory evoked responses recorded from post cardiac arrest patients in the intensive care unit, we studied separately standard and deviant responses (**Figure 1**), which is a novel and different paradigm compared to the classical MMN. We took into account the total 20 minutes extracted data, instead of the short interval response occurring in [100-300] ms following stimulation. We filtered the signal in the [0.5-50] Hz band. Finally,, all standard and deviant stimulations were averaged leading to a response in the time interval [0 - 1000] *ms*(standard) and [0 - 500] *ms* (deviant).

**Figure 1.**
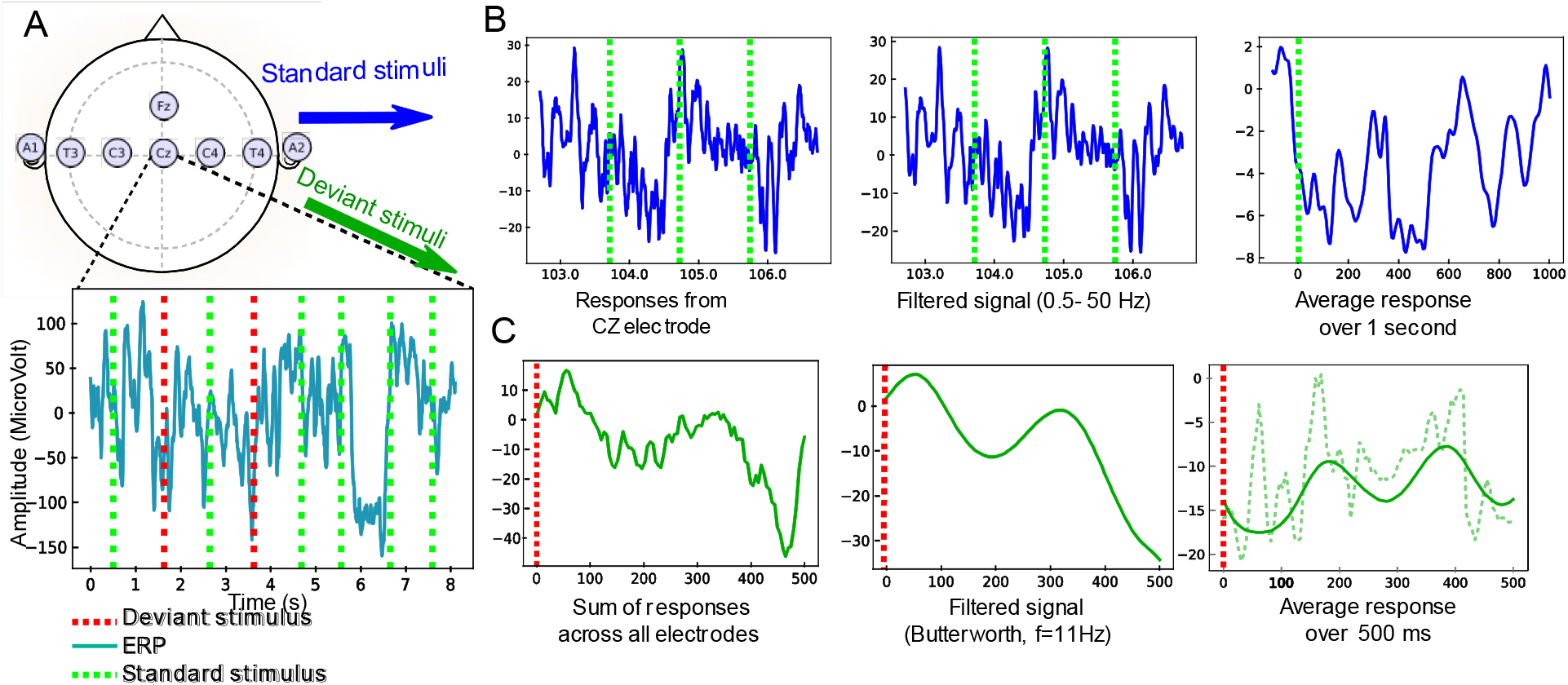
Pre-processing of the Evoked Auditory Responses to standard and deviant stimuli (an example of data obtained from the CZ electrode is given for standard stimuli and an example from all electrodes is given for deviant stimuli). **(A)** Upper: standard position of the EEG electrodes. Lower: EEG traces during a protocol mixing standard (green) and deviant (red) stimulations. **(B)** Sample of standard stimuli (blue) the EEG signal from CZ-electrode is filtered 0.5-50 Hz. The output is an average filtered response over one second. **(C)** Pre-processing of deviant stimuli: 1) the signal is summed over electrodes, 2) a low-pass filter is applied (Butterworth with ***n* = 2**, cutoff frequency at 10 Hz), 3) Average filtered response (continuous green) in a window of **500 *ms*** to a deviant stimulus, computed after synchronization to the stimulus. The non-filtered average response is also shown (dashed line).

We first focused on the ERP responses to standard periodic auditory stimuli, every 1s. We filtered the time series *X*(*t*) using a Butterworth bandpass filter (*n* = 4) in the frequency range 0.5-50Hz and obtained the output. Finally, we averaged the signal in the time interval [0 − 1]*s*, ensuring that auditory stimuli were produced at time *t* = *nT* (T=1s) leading to the response

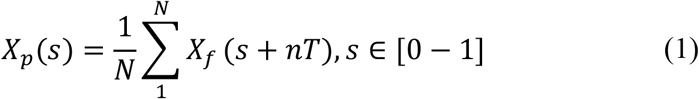

where *N* is the number of periods (typically of the order 10^3^). This preliminary procedure therefore allowed obtaining an average response *X*_*p*_ that highlights any possible deterministic feature present in the response. We applied a similar averaging procedure for deviant stimuli (see below and **Figure 1**).

##### 1. Analysis of responses to standard stimuli

For the analysis of standard stimulation, we divided the 20 minutes recording into 2 parts to compute temporal correlations. The main parameters we extracted to study the response to standard stimulations were defined as follows:

We computed the standard deviation *σ*_*X*_ of the signal in the time interval [20 − 320]*ms*: (as no synaptic integration is expected in less than 20ms. To note, analysis over the time window (20-500 ms) was not different from the one obtained over the 20-320 ms time window.

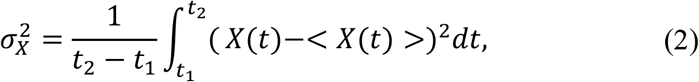

where *t*_2_ = 320*ms* and *t*_1_ = 20*ms*, and < *X*(*t*) > is the average of the X variable over the time [t1,t2]. This time interval corresponds to time scales of the neural networks involved in cognitive tasks.

We then divided the acquisition time of 20 minutes into n equal parts. For n=2, we got [1 − 10]*min* and [10 − 20]*min*. We averaged the signals on each of these periods to obtain two responses *X*(*t*) and *Y*(*t*) in the interval [0,1]*s*. We computed the time correlation or similarity in of these two signals:

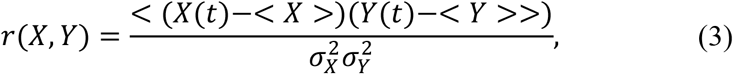

where <. >represents this time average is the average of the variable X.

We therefore used these two parameters to define the space state for the coordinates a patient: 1-the standard deviation computed over the entire sample of 20 min and 2-similarity, computed in equation 3. **). These coordinates define a mathematical state space, it is not a specific medical state of the patient**.

##### 2) Analysis of responses to deviant stimuli

Deviant stimuli are 14% of the entire responses. To define the signal of interest, we summed the signals from electrodes CZ, C3, C4 and FZ. We then filtered the resulting signal *X*_*d*_ using a lowpass Butterworth filter (n=2) with a cut off frequency at 10 Hz. Finally, we isolated responses in a window of 500 *ms* and computed average responses

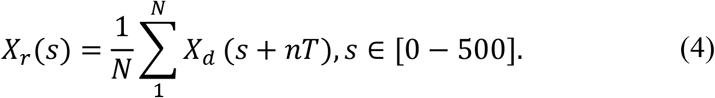

The smooth signal is shown in **Figure 1C**. We extracted two features:

1. The number *N*_*E*_ of local extrema (minima and maxima) in the response attained at points *e*_*i*_.
2. The total variation for the oscillation is measured by

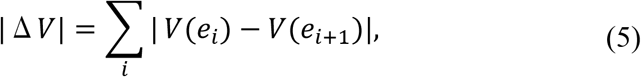

which is the sum of the absolute value of the difference between two consecutive extrema of the average evoked responses. This oscillation provides an information of the cumulative response amplitude.

#### 2- Features identification associated to standard and deviant responses

For standard responses, we subdivided the entire 20-min recording into two-time windows: the first 10 min (first period) and the period of the next 10 − 20 min (second period). We computed the variance (formula 2) and the correlation function (formula 3) of the response computed between the response in the first and second time period (**Figure 2A**). To test the ability of these two parameters to separate the two categories of patients, we plotted the histogram of these two parameters for all patients in our data (**Figure 2B**), showing that each parameter individually could be potentially used for a classification.

**Figure 2.**
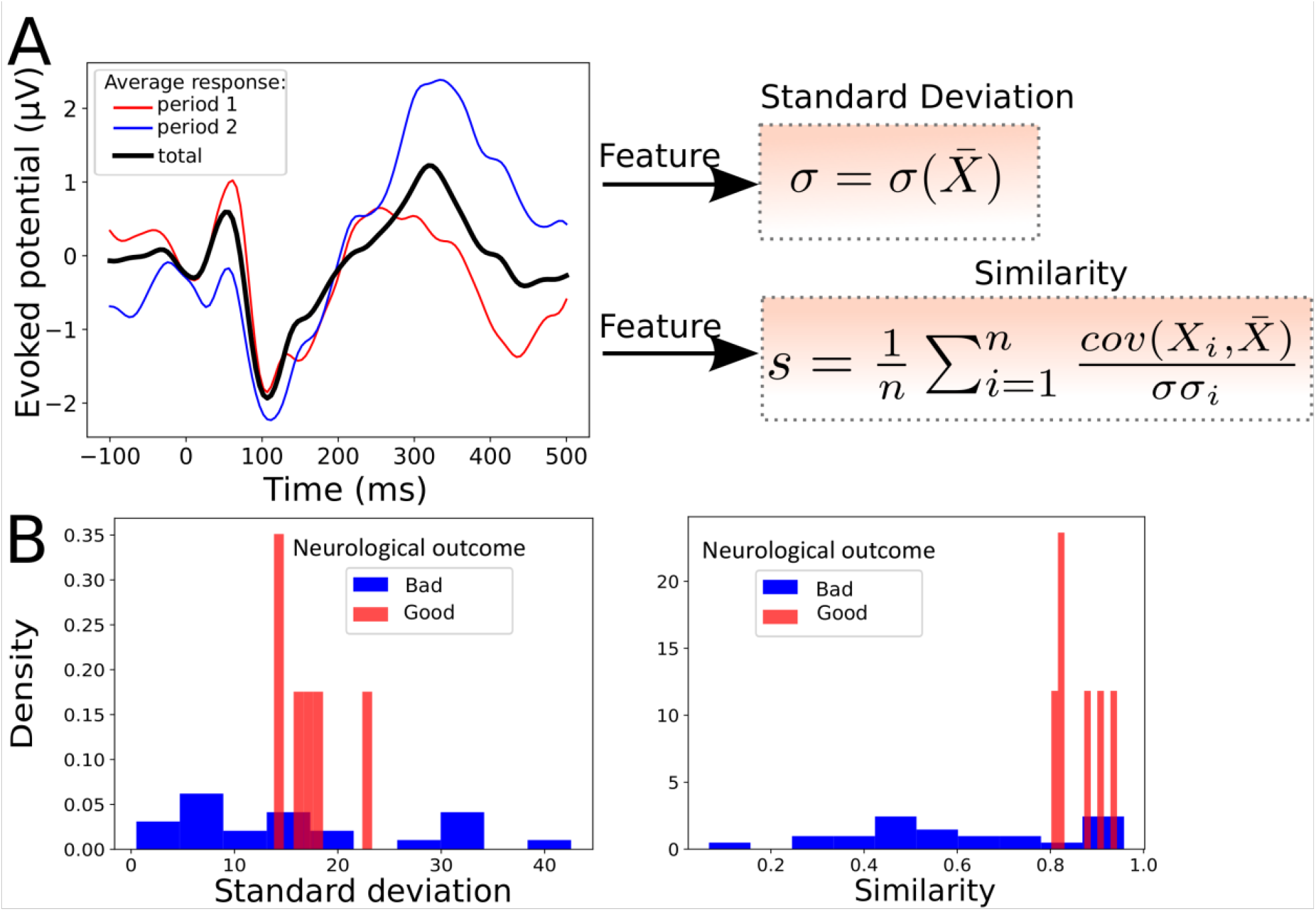
Statistical features associated to standard responses. **(A)** Left: Average evoked responses computed over a time window of **[0 − 10]*min*** (period 1, red), **[10 − 20]*min*** (period 2, blue) and over the entire period (**[0 − 20]*min***, black). Right: The standard deviation ***σ*** and the average correlation function ***s*** (similarity), between the response over the entire period (**[0 − 20]*min***) and over one of the n periods (**[0 − 20]*min*** or **[10 − 20]*min***), here **(B)**. Example of features distribution of dataset from the Cz electrode: standard deviation (Left) and similarity (Right) computed over the entire period; red (good neurological outcome) and blue (bad neurological outcome). These two parameters taken separately are insufficient to properly discriminate the two groups of patients.

For the deviant responses, as the signal showed different characteristics, we decided to use novel features: the first one consisted in the number of extremum *N*_*E*_ present in the signal (**Figure 3A**) and the second was the absolute value of the oscillation | Δ *V*|, which represents the sum of the differences between the extrema (formula 5). The result of this classification is shown by histograms of the two parameters computed over the whole population of patients (**Figure 3B**).

**Figure 3.**
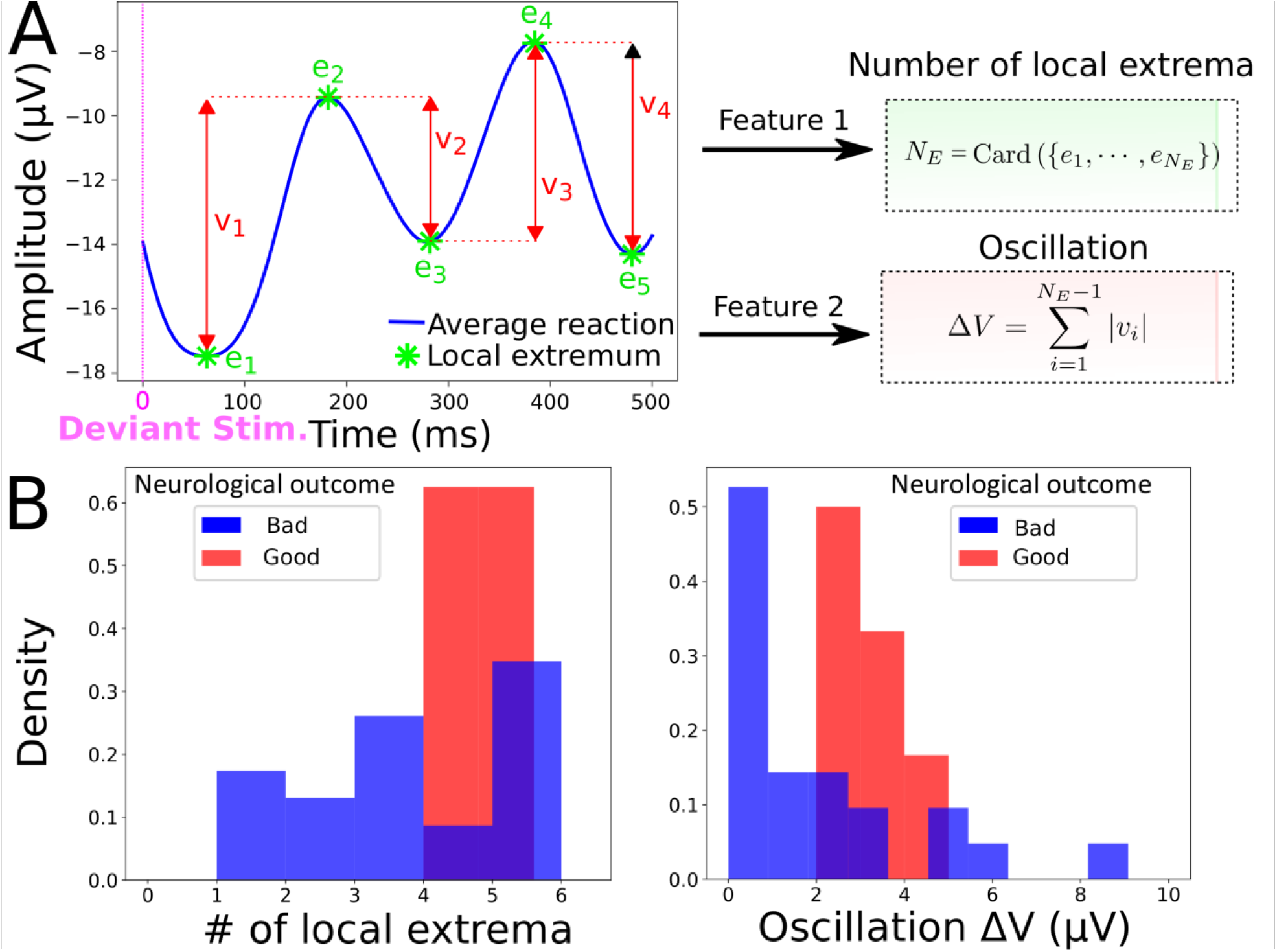
Statistical features associated to deviant responses. **(A)** Left: The average filtered evoked response (blue) to deviant stimuli computed over the entire time window contains ***N***_***E***_ local extrema ***e***_***i***_ (minimum or maximum), which is the first feature. The second feature is the oscillation | **Δ *V***| = **∑** | ***V*** (***e***_***i***_) **− *V***(***e***_***i*+1**_), which is the sum of the absolute value of the difference between two consecutive extrema of the average evoked response. **(B)** Example of feature distributions of dataset from all electrodes: local extrema (Left) and oscillation (Right) over the entire period; red (good neurological outcome) and blue (bad neurological outcome).

We selected two different types of parameters to study standard and deviant responses, but each of them taken individually was not sufficient to clearly separate the two categories of patients.

#### 3- Classification using a two-dimensional map

Based on the parameters we extracted in the previous subsection, we generated two-dimensional maps: for the map associated to standard stimuli, each patient has the *P* = (*σ*_*X*_,*r*(*X,Y*)) coordinates, while for deviant stimuli, we used the *P* = (*N*_*E*_, | Δ *V*|) coordinates. In various plots, we normalized the coordinates in a population (*X*_1_,..*X*_*n*_) by:

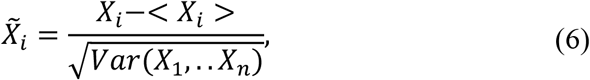

where < *X*_*i*_ > is the average over the points *X*_*i*_ and Var is the variance.

We mapped all points for all patients, where patients with bad *versus* good neurological outcome are shown in blue (vs red). Patients with good neurological outcome formed a cluster that will be the basis of the classification and prediction described below.

To study the maps defined above as predictive tools, we used three independent statistical classifiers (SVM, Gaussian estimator, K-nearest neighbours). We wished to assign a good neurological outcome probability to any point that would be added on the map based on the ensemble of previous data points already classified. Using the assumption that statistics associated to patients (features) are independent from each other, we used a Bayesian classification.

##### 1) SVM Classification

To classify the data, we used the standard SVM algorithm (Cherkassky and Mulier, 1998), which determines the hyperplane that best separates the two classes. Briefly, the chosen hyperplane maximizes the distance between itself and the closest points of each class, while all points of a given class are located on one of the two sides (Valiant, 1984). If no such hyperplane is found, which is the case here, the dimension of the space where the data are embedded is increased, a procedure known as kernelling (Aizerman AM, 1964). In a higher dimensional space, the classes are well separated by a higher dimensional hyperplane. If the two classes are still not well separated, a penalty is inflicted for every misclassified data point (Cortes and Vapnik, 1995). Here, the kernel is the Radial Basis Function *K*(*x,x*′) = exp (− *γ*| |*x* − *x*′ | |^2^), with and a penalty coefficient. Note that We obtained similar confusion matrix for all pairs (γ,C)∈[0.5,2.5]×[3,30] for SVM.

We implemented the SVM using the Scikit Learn module (Pedregosa et al., 2011; Buitinck *et al*., 2013). In general all our analysis and classification codes are written in homemade python script.

##### 2) Gaussian estimator

In case of a Gaussian estimator, we estimated the mean and the covariance matrix for the 2 categories of patients. The probability of each class is computed empirically using the maximum likelihood estimator (Supplementary methods). We recall that for an ensemble of n data 𝒮_*n*_ = (*x*_1_,..*x*_*n*_) that are separated into two classes, *C*_1_ and *C*_2_, the probability that a patient *X* belongs to one class, conditioned on the ensemble 𝒮_*n*_:

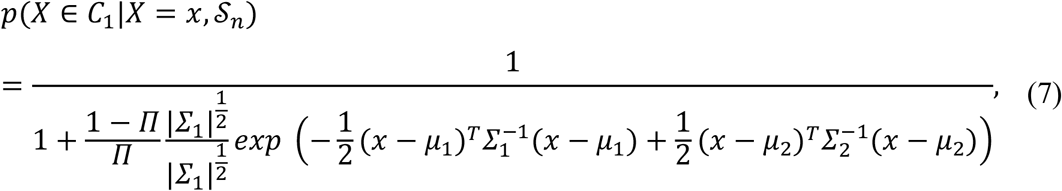

where (*μ*_*i*_, ∑_*i*_)_*i*=1,2_ are the mean and variance computed from each class *C*_1_ and *C*_2_ from 𝒮_*n*_. We used the fraction 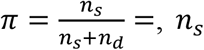 for the number of patients with good neurological outcome at discharge and *n*_*d*_ for the other patients. Formula 7 is derived in the supplementary methods.

##### 3) K-Nearest neighbours classifier and weighted K-Nearest neighbours

To classify the standard stimuli, we used the K-Nearest neighbours classifier. We computed the ratio for the probability of belonging to a class. For a given point *X*, the probability to belong to class *C*_1_ (“good neurological outcome”) given the distribution of point *X* is computed empirically as the number of neighbours out of a total of K.

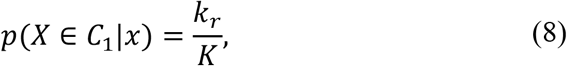

where *k*_*r*_ is the number of neighbours that belong to the class “good neurological outcome at discharge” among K closest points.

To classify deviant stimuli, we used a variant of the K-neighbours method by adding distance-relative weights to the points inside the dataset. The two classes labelled “bad neurological outcome” and “good neurological outcome” are defined as *C*_1_ and *C*_2_ respectively. The ensemble of points 𝒮_*n*_ in dimension 2 are given by the coordinates *x*_*n*_ = (*N*_*E,n*_, Δ *V*_*n*_), extracted in subsection “Analysis of responses to deviant stimuli”. To compute the classification probability, we defined K-nearest neighbourhood 𝒩_*K*_(*x*) for the point *x* as the *K* shortest points from *x*, computed from the Euclidean distance (between two points *x*_*n*_, *x*_*m*_),

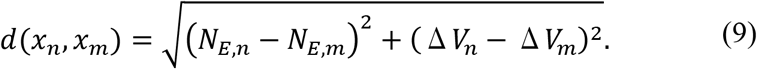

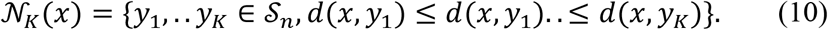

Finally, we used the weighted conditional probability defined by

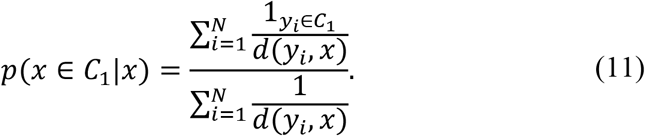

##### 4) Cross-validation

We used a cross-validation approach to validate the classification algorithm: we excluded a patient at a time and computed the probability of a good neurological outcome at discharge, based on the remaining elements in the data basis (Kohavi, 1995). We then computed this probability using the three classifiers, SVM, Gaussian estimators and K-neighbours and compared the result to the true result. We followed the protocol: 1- a patient *P*_*i*_,*i* = 1..*N* is selected inside the data basis; 2-we trained the classification algorithm on the database of all patients {*P*_*k*_.*k* = l..*N*} − *P*_*i*_. We evaluated the prediction of the model on the excluded patient, leading to a score *s*_*i*_. We recall that *s*_*i*_ = 1 if the prediction is correct, otherwise, *s*_*i*_ = 0. We then replaced the patient *P*_*i*_ inside the database and reiterated the procedure until each patient has been exactly excluded once. The final score of the model is computed as

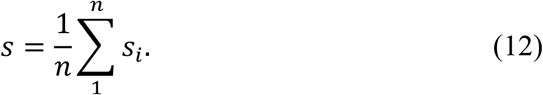

Finally, the confusion matrix defined as

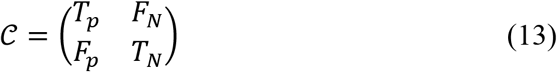

for the true positive *T*_*p*_ (number of patients who have a good neurological outcome at discharge and are classified correctly), true negative *T*_*n*_ (number of patients who have a bad neurological outcome and are classified correctly) and false positive *F*_*p*_ (number of patients who have a good neurological outcome and are classified incorrectly) and false negative *F*_*n*_ (number of patients who have a bad neurological outcome at discharge and are classified incorrectly). We calculated for each of the classifiers accuracy, sensitivity and specificity.

##### 5) Combined probability for outcome decision

We proposed to use for the predictive decisional probability *p*_*dec*_ the minimum of the ones estimated for the standard (relation 8) and deviant (relation 11) classifications. For a patient of coordinate *x* in each map, survival probability is:

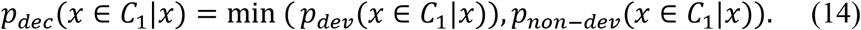

##### 6) Iteration and changing k-neighbours k

The approach developed here is iterative and any new additional case enriches the database and the classifications maps. For the K-neighbours approach, adding a point does not require any changes in the computation, although we expect that the number of neighbours that will enter progressively into the computation could diminish as the number of cases added in the map increases. For the Gaussian classification, the mean and the variance are recomputed following each new case.

## Results

### Overall Patient characteristics

Data of twenty-nine consecutive patients were analysed. Seven patients out of twenty-nine survived, but only 6 out of 7 were able to follow verbal command at hospital discharge. None of the patients was lost of follow-up but disability information was not available for one of them (back home without further information). At 3-6 months, GOS-E was scored at 3 for the patient who was unable to follow verbal command at discharge and died 27 months later without neurological improvement. GOS-E was scored at 4 for one patient, at 5 for one patient, at 6 for one patient and at 8 for the last two patients. Age, sex, medical history, characteristics of CA and electrophysiological features are presented in Table 1. At the time of recording, all patients were still hypothermic (< 35°C). Sedation was present in 11 out of the 29 patients (38%) at the moment of the electrophysiological recordings. For the non-surviving patients, 18 out of 22 died after withdrawal of life-sustaining therapies.

**Table 1.**
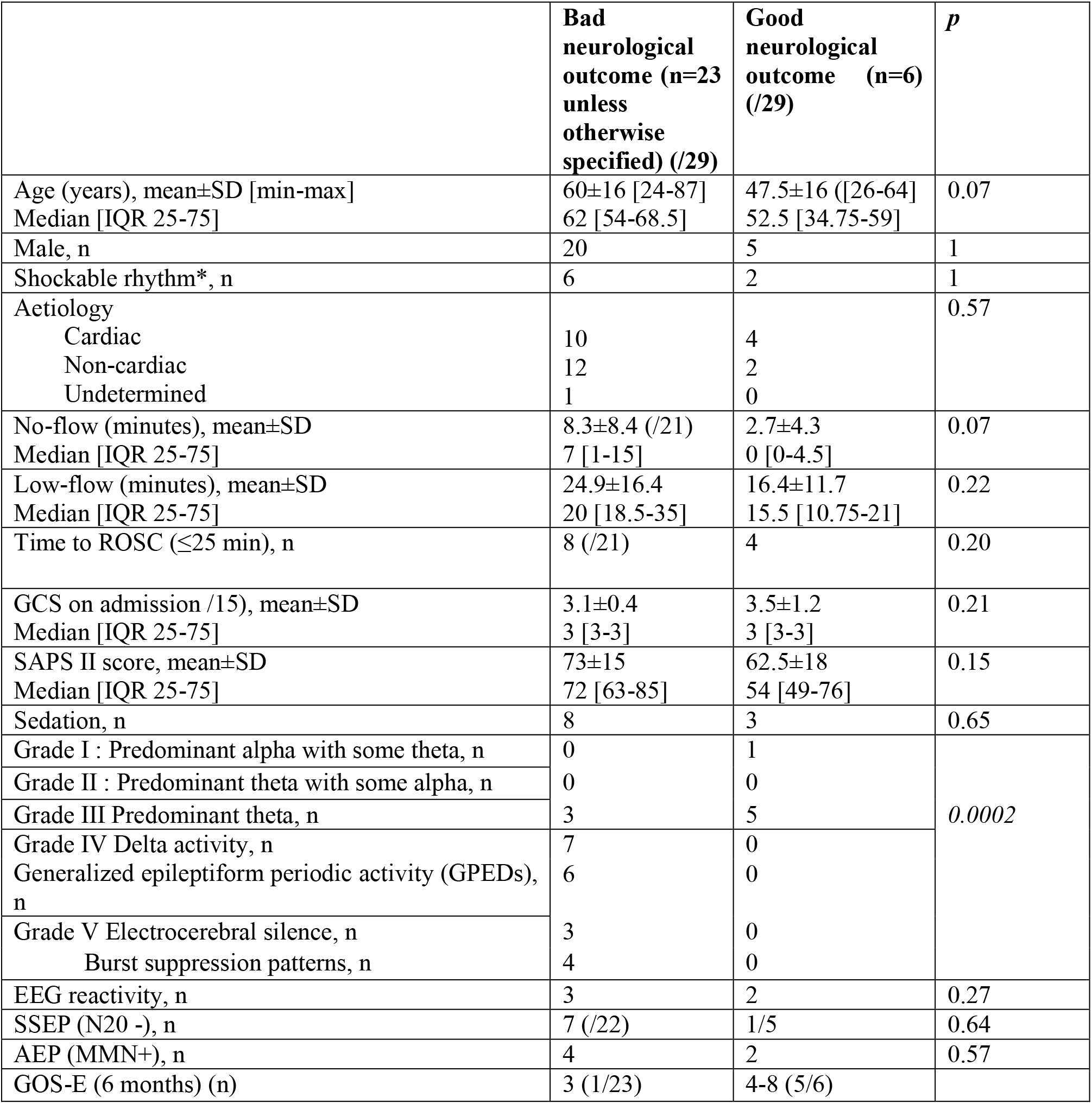
Comparison of clinical and electrophysiological characteristics between the two groups. *, as the first documented rhythm; ROSC, return of spontaneous circulation; GCS, Glasgow Coma Score; SAPS II score, simplified acute physiology score II; EEG, electroencephalography patterns according to the five major grades of severity scale for brain injury; SSEP, cortical somatosensory evoked potentials; AEP, auditory evoked potentials; MMN, mismatch negativity. No-flow data were missing in two patients and SSEP (N20 response) data in one patient (underlying Charcot Marie Tooth disease). GOS-E, Glasgow Outcome Scale-Extended.

The six patients with good neurological outcome presented an EEG pattern graded I to III for all, whereas 20 out of the 23 of the patients with final bad neurological outcome or death presented an EEG pattern graded IV or V (*p*<0.0002), including the patient who survived 27 months with bad neurological outcome. EEG reactivity (2/6 versus 3/23) and presence of MMN (2/6 versus 4/23) were more frequent in the group with good neurological outcome, whereas N20 was less frequently absent (1/5 versus 7/22), but none of these last markers were statistically different between the two groups. Only 2 patients presented congruent favourable prognostic factors with a present N20 at SSEP, a positive MMN and EEG pattern graded I to III (areactive EEG for both), among whom one patient did not survive. By contrast, four patients presented congruent bad prognosis factors with absent N20, absent MMN and an EEG pattern graded IV or V and all of them died. ERP obtained at Cz location were the most reproducible and the only ones used for visual analysis. Artefacts prevented the interpretation of SSEP in one patient of each group.

### Prognosis map constructed from Bayesian statistical inference

Since each parameter taken individually for standard (standard deviation and similarity) and deviant (number of extremum *N*_*E*_ and oscillation | Δ *V*|) responses were not sufficient to obtain a clear separation between the two patient categories, we decided to combine them into a two-dimensional map (**Figure 4**). Interestingly, we found that this map allowed a clear separation that we quantified using various *a priori* classifiers: SVM, Gaussian and the K-neighbour classifiers (Hastie et al., 2001) (Table 2).

**Table 2.** Accuracy, sensitivity and specificity obtained by cross-validation for responses to standard, deviant stimuli; k is the number of neighbours used in the classification algorithm.

|  | k=3 | k=4 | k=6 | k=8 | SVM | Gaussian |
| --- | --- | --- | --- | --- | --- | --- |
| **Accuracy** |  |  |  |  |  |  |
| Standard Responses | 0.96 | 0.96 | 0.89 | 0.85 | 1.0 | 0.92 |
| Deviant Responses | 0.89 | 0.89 | 0.89 | 0.89 | 1.0 | 0.89 |
| **Sensitivity** |  |  |  |  |  |  |
| Standard Responses | 0.83 | 0.83 | 0.83 | 0.66 | 1.0 | 0.66 |
| Deviant Responses | 0.83 | 0.83 | 0.83 | 0.83 | 1.0 | 0.5 |
| **Specificity** |  |  |  |  |  |  |
| Standard Responses | 1.0 | 1.0 | 0.9 | 0.9 | 1.0 | 1.0 |
| Deviant Responses | 0.91 | 0.91 | 0.91 | 0.91 | 1.0 | 1.0 |

**Figure 4.**
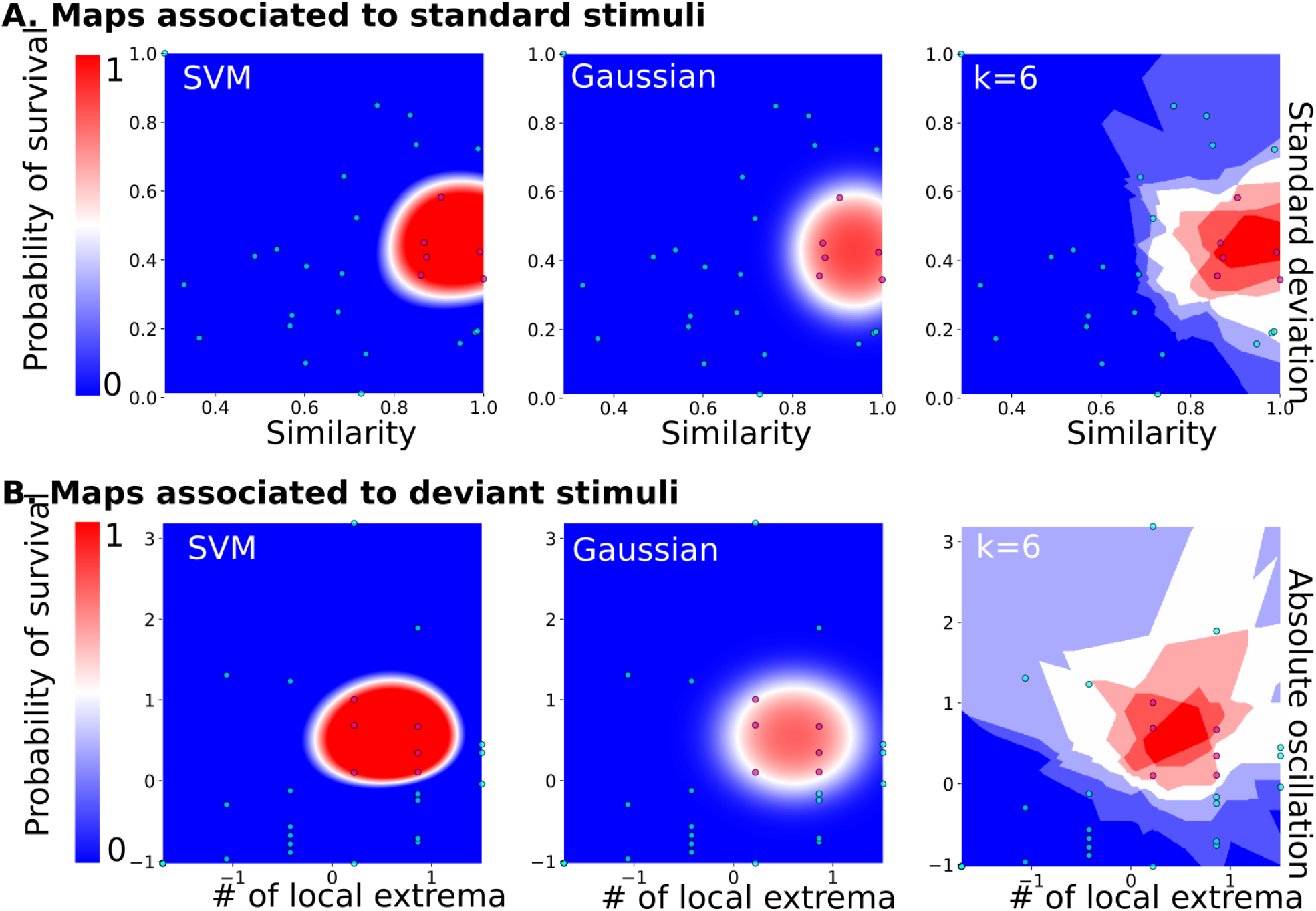
Predictive probability maps of good neurological outcome. **(A)** Probability maps computed from features of the standard stimuli responses. From left to right: maps computed from SVM, Gaussian and the k-Nearest Neighbours classifier (k=6, the worst case scenario). **(B)** Probability maps computed from features of the deviant stimuli features. From Left to right: maps computed from SVM, Gaussian and the k-Nearest Neighbours classifier with distance-related weights, k=6 for example.

The classification probability of a patient characterized by its coordinates was obtained by computing a score that measures the proximity to one of the two categories of patients.

When mapping all the features first taken individually for standard and deviant responses, we found a cluster formed of patients with a “good” neurological outcome, bounded in red, well separated from the area in which were found the other patients (non-surviving or “bad” neurological outcome). This partition between two distinct areas was present in all classifiers: SVM, Gaussian and k-neighbours, confirming that this partition was robust independently of the choice of the classification methods (supplementary **Figure S1** for other choices of k for the k-neighbour algorithm). We found a similar partition into two categories of patients when classifying the standard or the deviant responses (**Figure 4**). Because the present database did not contain many patients and to guarantee the robustness of our approach, we decided to use three classifiers (SVM, Gaussian mixture and k-neighbours) that ultimately converged and gave similar results. As a small size database is also associated with overlearning, and to overcome this difficulty, we chose to use simple models for classification: Support Vector Machines (SVM) seems to be particularly suitable, as its classification was dependent only on a reduced number of patients.

To obtain an accurate classifier, we used a different version of the K-neighbours classification, where the weight depends on the distance between the point to classify and the K-nearest neighbours (formula 11). The present classification maps for both standard and deviant responses studied separately showed that the neurological outcome of post-anoxic comas can be predicted (Table 2). Combining the probability computed in each map, we proposed a decision probability with a high specificity, which does not misclassify patients with good neurological outcome in the category of patients with bad neurological outcome.

### Classification efficiency of the two-dimensional maps

To test the predictive strength of the standard and deviant responses classification, we computed the confusion matrix (formula 13) as described in the methods. The confusion matrix computed for Gaussian estimator showed a 89 % accuracy, and a 100% validation accuracy for SVM. The confusion matrix computed for k-neighbours was less performing than SVM. The sensitivity remains high and could be improved with the increasing number of classified patients (k=4; similar results were obtained for other values of k) with distance-dependent weight showed this estimator introduces type I error, with an accuracy of 0.90, a sensitivity of 0.83 and a specificity of 0.91.

Finally, we also computed the confusion matrix obtained from MMN obtained by visual analysis, we obtain an accuracy of 0.72, a sensitivity of 0.33 and a specificity of 0.82. If MMN remains an interesting indicator it shows a very weak sensitivity in these patients (Tables 2, 3).

**Table 3.**
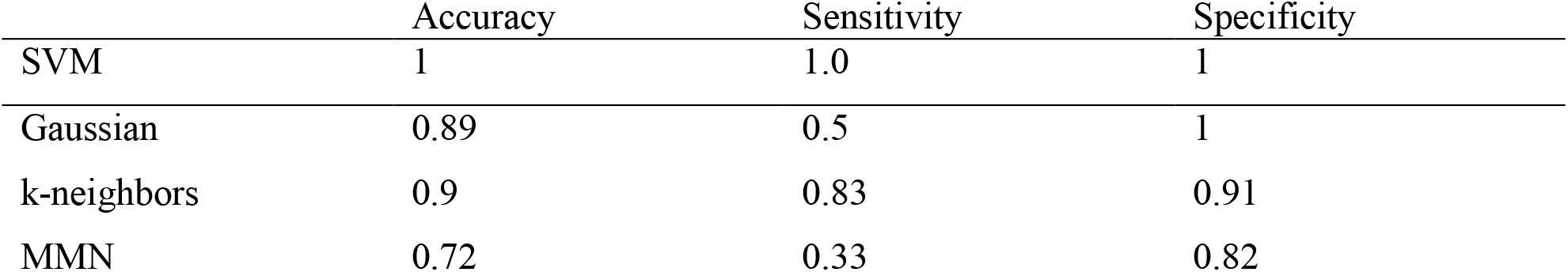
Classifications scores

|  | Accuracy | Sensitivity | Specificity |
| --- | --- | --- | --- |
| SVM | 1 | 1.0 | 1 |
| Gaussian | 0.89 | 0.5 | 1 |
| k-neighbors | 0.9 | 0.83 | 0.91 |
| MMN | 0.72 | 0.33 | 0.82 |

## Discussion

Our exploratory study was designed to identify new features from the AEP recording that could be more powerful than visual inspection of MMN in the routine ICU setting. The originality of the present strategy was to consider independently deviant from standard responses and to identify new features from the total amount of information that is generated during the procedure. Very importantly, we did not select them to obtain a best separation of patients, but it turned out that we obtained a good separation using this step-by-step analysis described in the method’s section. The output of this novel classification method is good neurological outcome probability and allows clustering patients in one of the two categories, within a few seconds without a specialist interpretation.

Characterizing electrophysiological features to predict the outcome of post-anoxic coma remains a genuine challenge. There is currently no satisfactory, efficient and simple tool to predict comatose patient outcome accurately, especially at the acute phase, when patients are sedated and/or hypothermic. Standard electroencephalography is the most common method used to predict prognosis in those patients. If highly malignant pattern (suppressed background discharges without discharges or with continuous periodic discharges, or burst suppression background with or without discharges) is highly specific of poor outcome, as shown in our study, it has a sensitivity of only 50% (Westhall et al., 2016). The absence of cortical N20 response at SSEP after stimulation of median nerves has an almost 100% specificity for non-awakening prediction (Sandroni et al., 2014), whereas the predictive value of the visual analysis of MMN for post-CA comatose patients, limited to a binary response (presence/absence of a detectable peak of the MMN between the standard and deviant responses) is poorly sensitive (Azabou et al., 2018), as shown in our study. Several statistical methods have been developed to improve accuracy, some based on sample-by-sample paired t-test in a specific time window, while others based on wavelet transform, multivariate, cross-correlation and probabilistic methods (Fischer et al., 1999) (Gabriel et al., 2016) (De Lucia and Tzovara, 2016) (Naccache et al., 2005) (Daltrozzo et al., 2007) (De Lucia and Tzovara, 2015) (Naccache et al., 2015) (Juan et al., 2016). Overall, this explains why a multimodal prognostication approach is still recommended in these patients including clinical examination and serum biomarkers (Sandroni et al., 2014). Here, we present a novel approach that considered the amount of information not analysed by the classical interpretation of MMN, which uses only the subtraction of the standard and deviant responses. We analyzed them first separately, then in combination, to evaluate the additive value of each in the prognosis assessment. On one hand, we analyzed responses to standard auditory stimulations using variance and similarity. On the other hand, we analyzed responses to deviant auditory stimulations using two novel parameters (number of extrema and total length of the response between these extrema), which measure the cumulated oscillation of the ERP signal over hundreds of ms. After extracting features from the time average responses computed during the first 500 *ms*, we constructed predictive maps in two dimensions. To evaluate the robustness of our method, we used three classifiers, showing similar maps classification results. Interestingly, the area of patients with good neurological outcome formed a well-bounded cluster, demonstrating that these patients had comparable and similar EEG features. Finally, using cross-validation statistics, we computed a score of classifications, demonstrating that any of the three classification methods was more robust than simply analysing the Mismatch negativity in a binary response, using logistic regression or single-trial topographic analysis (De Lucia and Tzovara, 2015). At last, we validated this approach based on four electrodes with one single electrode Cz. We showed that good neurological prognosis probability maps allow us to predict the neurological outcome of post-anoxic comatose patients with a very good accuracy of 0.90, sensitivity of 0.83 and specificity of 0.91 when considering the least efficient classifier (Tables 2 and0 3).

The two classes of auditory stimulations are neither redundant nor fully correlated and we used different features to generate classification maps. In the present study, we analysed a single data set that lead to a classification map. In order not to classify a patient who will have a good prognosis outcome wrongly, i.e. with a bad neurological outcome, we choose among several classifiers the one that had the highest specificity. We performed here a cross validation analysis where we separated the patient database into a testing and a training group. We divided the database into one group with 1 patient out and a second group with the other 28 patients. We ran this test 28 times so that each of the 29 patients was alternatively included in the 1 group patient and this allowed us to reclassify with a given probability for each patient outcome based on the new map determined by the other patients. The robustness resulted from the comparable results for each of standard and deviant auditory stimulations considered independently (**Figure. 4**): we obtained convergent results, which is a satisfactory confirmation of the predictions. Finally, as commonly done in statistical classification, we increased the dimension of the parameter space to better separate the data points. By using two variables, it was sufficient to represent the data into a space where the separation between “bad” and “good outcome” was possible. The robustness of the present approach was confirmed by the fact that we obtained comparable results for each of standard and deviant auditory stimulations considered independently using three different classifiers (**Figure 4, Tables 2 and 3**) and at this stage, this was the best we could do to assess the robustness of our method.

Tzovara et al. already reported that the progression of MMN auditory discrimination over the first two days of coma is of good prognosis (Tzovara et al., 2013), suggesting that collecting repetitive data within days, or at an earlier phase, could reveal changes that could have a higher predictive value. In view of a large dissemination, and besides the potential additive value of repeating examinations, it would also be interesting to test if this procedure is generalizable to other auditory oddball paradigms. In that respect, analyzing EEG in which diverse auditory and nociceptive stimuli are applied during recording would also be interesting.

The physiopathological significance of our findings remains speculative. Conscious states require a brain scale communication and does not rely on a single neuronal network. Excitatory neurotransmission is impaired across cortico-cortical, thalamo-cortical and thalamo-striatal connections (Edlow et al., 2021). Auditory discrimination is particularly sensitive to fluctuations in the state of consciousness (Górska et al., 2021), (Wu et al., 2018). Responses in those studies have been reported to be maximal at frontocentral locations (Fz and Cz), which are also electrodes where connectivity studies have shown the maximal response (Binder et al., 2017). The total amount of data we collected for all epochs during the 20 minutes of auditory stimulations and not only during the time window used for MMN might explain our more sensitive results. However, the present method do not necessarily reflect specific cognitive processes, and it is pre-mature at this stage to relate the chosen metrics to underlying brain injury or cognitive processes.

In that small series, none of the classical electrophysiological tools were sensitive or specific enough to give a reliable neurological prognosis. Only 2 patients presented congruent favourable prognostic factors with a present N20 at SSEP, a positive MMN and EEG pattern graded I to III (benign pattern according to the ACNS EEG terminology) (Westhall et al., 2015) and areactive for both, among whom one patient did not survive. By contrast, four patients presented congruent bad prognosis factors with absent N20, absent MMN and an EEG pattern graded IV or V (highly malignant pattern according to the ACNS EEG terminology) and all of them died, suggesting that congruent pejorative factors are strongest indicators of prognosis than congruent good prognosis factors, in accordance with literature. It is to note that one third of the patients with bad outcome and 50% of the patients with good outcome were under sedation at the time of recording, which is known to impede electrophysiology interpretation. Our study was not designed to compare our tool with classical electrophysiological examinations but sensitivity and specificity were higher in that small cohort that needs to be validated in a larger cohort.

That pioneer study has several limitations. First, as a retrospective study, neurological prognosis was evaluated on the ability of the patient to follow verbal command at discharge, which remains a subjective assessment that may have led to patient’s misclassification. In the 7 surviving patients, GOS-E was available for 6 of them at 3 to 6 months post-discharge and was found at 3 in the patient who was initially unable to follow verbal command and from 4 to 8 for the others. Because of the retrospective design of our study, withdrawal of life-sustaining therapies decisions had been taken before our new analysis. They were multimodal and based upon European guidelines ERC-ESICM (2014). Second, this cohort may not be representative of all post CA patients since electrophysiological assessment was performed relatively late, up to 6 days after admission, in patients still comatose at the time of the evaluation. Third, the relatively small sample size prevents generalization of our results. Fourth, we tried to offset the imbalanced cohort between patients with good and bad prognosis using a leave-one out cross validation with a testing and training group, and used three different classifiers to increase the robustness of our method. Fifth, our new approach did not consider the order of the different sounds. For instance, a standard sound that would start a new sequence just after a deviant sound or ending a series of standard sounds just before a deviant sound, may not be processed the same. This point could deserve a specific attention in future studies, but as we averaged all our data, this probably does not bias our results. At last, even if the study was not designed to compare the classical neurophysiological approach to this algorithm, each examination had insufficient specificity and sensitivity, in particular concerning MMN. The choice of the MMN paradigm we used could be criticized. However, visual analysis of MMN is not robust enough to be reliable, even when choosing parameters that better discriminate standard and deviant sounds (Azabou et al., 2018).

To conclude, we developed a new promising classification method that could be self-sufficient, easily used by intensivists (only one electrode, with minimal cost and easy training), without the help of the neurophysiologists and in sedated and/or hypothermic patients, since these conditions represent actual limitations to electrophysiological data acquisition in the ICU. The produced maps can be refined and upgraded by adding new cases and thus increase the performance of the probabilistic classifier. In the future, and according to the local human and logistical resources, the software could be implemented with other electrophysiological and clinical variables to provide an optimal estimated probability of the patient outcome, independently from neurophysiologists. Developing such algorithms, ready-to-use by the intensivits, would enable more aggressive management in patients with predicted good neurological outcome and conversely, avoid usefulness interventions in those with predicted bad neurological outcome. Whether this approach could be secondarily applied to other predictive situations and generalized to other comas remains to be validated.

## Data Availability

All data produced in the present study are available upon reasonable request to the authors

## Declarations

### Ethic approval and consent to participate

The study was approved by the ethics committee (Comité de Protection des Personnes, CPP Paris IV #2012/22) (PHRC CAPACITY AOR10109).

### Consent for publication

Information was provided to next-of-kin for all participants, followed whenever possible, by information to the patient.

### Data availability

The datasets generated during and/or analysed during the current study are available from the corresponding author on reasonable request.

### Competiting interests

FA, AR, NK and DH have a patent application for the prediction of coma outcome (French patent FR1852473, titled “Outil prédictif de la sortie du coma des patients après un arrêt cardio-respiratoire”).

### Author contributions

AF, AD, DH created the algorithm. SL, ND and BM collected the clinical data. AR, EM, SM, PL and NK performed the electrophysiological examinations. DH and NK wrote the manuscript. All authors read and approved the final manuscript.

## Acknowledgements

Not applicable

---

**Figure SI1.**
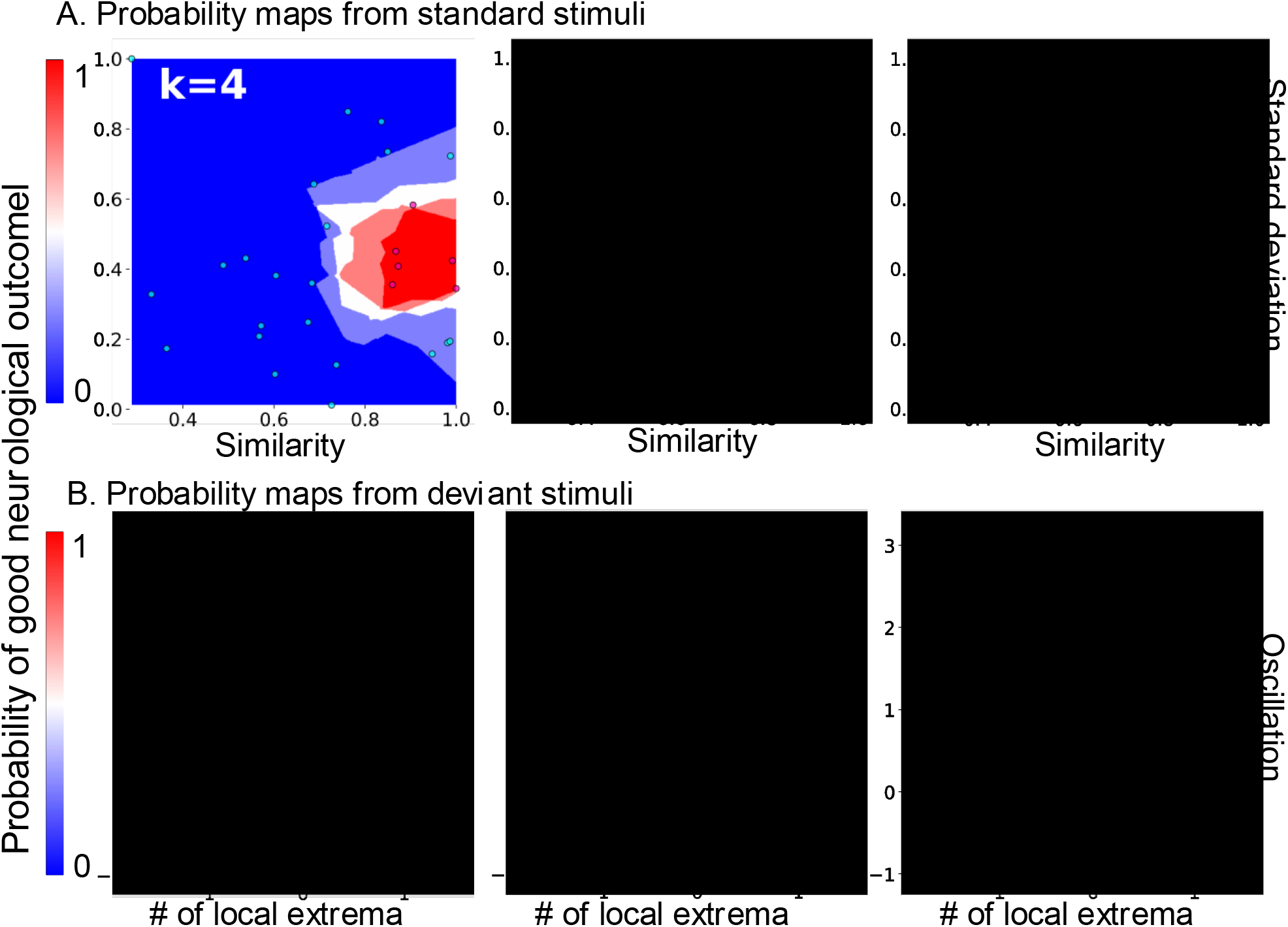

