## Supplementary material for "Two-dimensional maps to predict the neurological recovery after cardiac arrest": SI

### Supplementary methods

#### Computing the A priori estimators

We compute here the a priori Gaussian empirical estimators. In the framework of two classes of patients with good versus bad neurological outcome that we consider to be normally multidimensional distributed, we will first derive the estimator for a point $x$ to be classified, based on the mean and variance, that we relate to the sample of the database of patients with good neurological outcome. The computations use the Bayes'rule and we assume that each class can be distinguished by their mean and covariance matrices, which should be a priori different. The variable $y$ represents the classification on the class, while x represents the position in the phase space. We assume the following apriori probability:

$$y \sim \text{Bernoulli}\,(\Pi), \quad x|y \sim \text{Normale}\,.(\mu_i, \Sigma_i) \qquad (29)$$

The parameter to be estimated are $\theta = (\Pi, \mu_0, \mu_1, \Sigma_0, \Sigma_1)$. The data base $\mathcal{S}_n$ is of size $n$. The log-likelihood estimator is

$$\begin{aligned} l(\theta) &= \log\left(\prod_{i=1}^{n} p\,(x_i, y_i|\theta)\right) = \sum_{i=1}^{n} \log\,(p(x_i, y_i|\theta)) \\ &= \sum_{i=1}^{n} \log\,(p(x_i|y_i,\theta)p(y_i|\theta)) \end{aligned} \qquad (30)$$

Splitting the sum with respect to the two classes represented by $y = 1$ for $(p(y|\theta) = \Pi)$ and $y = 0$ $(p(y|\theta) = 1 - \Pi)$, we get:

$$\begin{aligned} l(\theta) &= \sum_{y_i=1} \log\,(p(x_i|y_i,\theta)p(y_i|\theta)) \\ &+ \sum_{y_i=0} \log\,(p(x_i|y_i,\theta)p(y_i|\theta)) \end{aligned} \qquad (31)$$

Then:

$$\begin{aligned} l(\theta) &= \sum_{x_i, y_i=1} \log\left(\Pi\,\frac{e^{\frac{-1}{2}(x_i-\mu_1)^T \Sigma_1^{-1}(x_i-\mu_1)}}{2\pi|\Sigma_1|^{\frac{1}{2}}}\right) \\ &+ \sum_{x_i, y_i=0} \log\left((1-\Pi)\,\frac{e^{\frac{-1}{2}(x_i-\mu_0)^T \Sigma_0^{-1}(x_i-\mu_0)}}{2\pi|a_0|^{\frac{1}{2}}}\right) \end{aligned} \qquad (32)$$

The total number of points in class 1 is

$$N = \sum_{y_i=1} 1. \tag{33}$$

We finally get:

$$l(\theta) = N\log(\Pi) + (n-N)\log(1-\Pi) - N\log(2\pi|\Sigma_1|^{\frac{1}{2}}) - (n-N)\log(2\pi|\Sigma_0|^{\frac{1}{2}}) - \frac{1}{2}\sum_{y_i=1}(x_i-\mu_1)^T\Sigma_1^{-1}(x_i-\mu_1) - \frac{1}{2}\sum_{y_i=0}(x_i-\mu_0)^T\Sigma_0^{-1}(x_i - \mu_0) \tag{34}$$

The value of the different parameter are obtained at the extremum of the estimators. Thus,

$$0 = \frac{dl(\theta)}{d\Pi} = \frac{N}{\Pi} - \frac{n-N}{1-\Pi} \tag{35}$$

leading to the empirical estimator

$$\widehat{\Pi} = \frac{N}{n}. \tag{36}$$

Differentiating with the mean, we get:

$$0 = \frac{dl(\theta)}{d\mu_0} = \sum_{y_i=0}(-\Sigma^{-1}\mu_0 + \Sigma^{-1}x_i) \tag{37}$$

Thus

$$\hat{\mu}_0 = \frac{\sum_{y_i=0} x_i}{n-N} \tag{38}$$

Similarly,

$$\hat{\mu}_1 = \frac{\sum_{y_i=1} x_i}{N}. \tag{39}$$

Finally,

$$\frac{dl(\theta)}{d\Sigma_1^{-1}} = \frac{N}{2}\frac{d(|\Sigma_1^{-1}|)}{d\Sigma_1^{-1}} - \frac{1}{2}\frac{d(Trace(\Sigma_1^{-1}\sum_{y_i=1}(x_i-\mu_1)^T(x_i-\mu_1)))}{d\Sigma_1^{-1}}, \tag{40}$$

and we recall that $\frac{d(log\,(|A|))}{dA} = A^{-T}$ et $\frac{d(Trace(AB))}{dA} = B^T$, indeed:

$$\frac{\partial}{\partial X_{ij}}\log(|X|) = \frac{1}{|X|}\frac{\partial |X|}{\partial X_{ij}} = \frac{1}{|X|} comX_{ij} = X_{ij}^{-T} \tag{41}$$

Using that $(comA)^T = |A|A^{-1}$ for an invertible matrix, we get

$$\frac{\partial}{\partial X_{ij}} Trace(XB) = \frac{\partial}{\partial X_{ij}}\sum_l \sum_k X_{lk} B_{kl} = B_{ji} = B_{ij}^T. \tag{42}$$

Then,

$$\frac{dl(\theta)}{d\Sigma_1^{-1}} = \frac{N}{2}\Sigma_1^{-T} - \frac{1}{2}\sum_i (x_i - \mu_1)^T(x_i - \mu_1) = 0 \tag{43}$$

$$\widehat{\Sigma_1} = \frac{1}{N}\sum_{y_i=1} (x_i - \widehat{\mu_1})(x_i - \widehat{\mu_1})^T \tag{44}$$

Similarly

$$\widehat{\Sigma_0} = \frac{1}{n-N}\sum_{y_i=0} (x_i - \widehat{\mu_0})(x_i - \widehat{\mu_0})^T. \tag{45}$$

We conclude with the final a priori probability:

$$\begin{aligned} p(y=1|x) &= \frac{p(x|y=1)p(y=1)}{p(x)} \\ &= \frac{p(x|y=1)p(y=1)}{p(x|y=1)p(y=1)+p(x|y=0)p(y=0)} \\ &= \frac{1}{1+\frac{p(x|y=0)p(y=0)}{p(x|y=1)p(y=1)}} \end{aligned} \tag{46}$$

$$p(X \in C_1|X=x) = \frac{1}{1+\frac{1-\Pi}{\Pi}\frac{|\Sigma_1|^{\frac{1}{2}}}{|\Sigma_0|^{\frac{1}{2}}} exp\left(-\frac{1}{2}(x-\mu_0)^T\Sigma_0^{-1}(x-\mu_0)+\frac{1}{2}(x-\mu_1)^T\Sigma_1^{-1}(x-\mu_1)\right)} \tag{47}$$

We use relation 47 to estimate the probability for a point $x$ to below to a given class, after the parameters are estimated from the ensemble of data.

#### Supplementary figures legends

**Supplementary Figure S1. K-neighbor classification maps for $\boldsymbol{k = 4, 6}$ and 8. (A)** Probability maps computed with features extracted from the standard auditory stimulations (Similarity and variance). **(B)** Probability maps computed from features extracted from deviant auditory stimulations (number of local extrema and oscillation). The red cluster does not disrupted despite the increasing k from 4 to 8, $\boldsymbol{k = 4}$ remaining the most robust value.
